# *Streptococcus pneumoniae* serotype 3 population structure in the era of conjugate vaccines, 2001-2018

**DOI:** 10.1101/2023.08.29.23294772

**Authors:** Eleonora Cella, Catherine G Sutcliffe, Lindsay R Grant, Carol Tso, Robert C Weatherholtz, Shea Littlepage, Ladonna Becenti, Mohammad Jubair, Brenna C Simons, Marcella Harker-Jones, Raymond Reid, Del Yazzie, Mathuram Santosham, Katherine L O’Brien, Laura L Hammitt, Taj Azarian

**Author notes:** Current affiliations: LRG – Pfizer; MJ- icddr,b; KLOB – World Health Organization. Contributed equally (Laura L Hammitt, Taj Azarian). Corresponding author: Laura Hammitt, 415 N. Washington St., Baltimore, MD 21231.

## Abstract

**Background:** Despite use of highly effective conjugate vaccines, invasive pneumococcal disease (IPD) remains a leading cause of morbidity and mortality and disproportionately affects Indigenous populations. Although included in the 13-valent pneumococcal conjugate vaccine (PCV13), which was introduced in 2010, serotype 3 (ST3) continues to cause disease among Indigenous communities in the Southwest US. In the Navajo Nation, ST3 IPD incidence increased among adults (3.8/100,000 in 2001-2009 and 6.2/100,000 in 2011-2019); in children the disease persisted although the rates dropped from 5.8/100,000 to 2.3/100,000.

**Methods:** We analyzed the genomic epidemiology of ST3 isolates collected from 129 adults and 63 children with pneumococcal carriage (n=61) or IPD (n=131) from 2001–2018 of the Navajo Nation. Using whole-genome sequencing data, we determined clade membership and assessed changes in ST3 population structure over time.

**Results:** The ST3 population structure was characterized by three dominant subpopulations: *clade II* (n=90, 46.9%) and *clade Iα* (n=59, 30.7%), which fall into Clonal Complex (CC) 180, and a non-CC180 clade (n=43, 22.4%). The proportion of *clade II*-associated IPD cases increased significantly from 2001-2010 to 2011-2018 among adults (23.1% to 71.8%; p<0.001) but not in children (27.3% to 33.3%; p=0.84). Over the same period, the proportion of *clade II-* associated carriage increased; this was statistically significant among children (23.3% to 52.6%; p=0.04) but not adults (0% to 50.0%, p=0.08).

**Conclusions:** In this setting with persistent ST3 IPD and carriage, *clade II* has increased since 2010. Genomic changes may be contributing to the observed trends in ST3 carriage and disease over time.

## Introduction

*Streptococcus pneumoniae* is gram-positive and extracellular colonizer of the human upper respiratory tract that can cause a range of infections including otitis media, bacteremia, pneumonia, and meningitis [1]. The capsular polysaccharide of the pneumococcus defines its serotype and is the main virulence determinant as well as the target for pneumococcal conjugate vaccines (PCV) [2]. As of 2020, over one hundred serotypes have been identified [3,4]. The carriage prevalence of *S. pneumoniae* in the nasopharynx varies with age, environment, and the presence of upper respiratory infections and, when assessed by culture methods, typically ranges from 5-10% among adults without children and 20-60% in school-age children [2]. Rates and duration of carriage also vary by serotype [5] and population characteristics. Despite effective conjugate vaccines, pneumococcal disease remains a significant cause of morbidity and mortality [6–8]. The case fatality rate of invasive pneumococcal disease (IPD) may reach 15-20% in adults and 30-40% in the elderly in high income settings [9–11].

While *S. pneumoniae* serotype 3 is included in the 13-valent pneumococcal conjugate vaccine (PCV13), observational studies of IPD in children and community-associated pneumoniae in adults have demonstrated varying effectiveness against this serotype [12,13]. Recent studies have found that the incidence of serotype 3 IPD has been increasing [14,15], and in the United States, serotype 3 was the most common PCV13 serotype identified in cases of IPD in 2017-2018 among adults [16]. We previously showed this increase is partially attributed to an emerging sub-lineage, termed *clade II* [17], belonging to Clonal Complex (CC) 180, which is the most common sequence type of serotype 3 worldwide. Through detailed genomic analysis of a global sample, *clade II* was found to possess antibiotic resistance and diverse surface protein antigens as compared to the prototypical *clade I* strains. However, phenotypic analysis did not indicate differences in capsule shedding or capsule charge, which have both been associated with differences in virulence, or susceptibility to antisera [17]. A subsequent analysis of serotype 3 IPD cases in England and Wales also demonstrated a significant increase in *clade II* after 2013 [18], with similar results in South America [19]. Yet, the underlying dynamics of the change in population structure associated with the emergence of *clade II* and the resulting impact on IPD rates remains unclear.

PCV7, introduced in 2010 for routine childhood immunization in the US, was designed to protect against pneumococcal serotypes 4, 6B, 9V, 14, 18C, 19F and 23F. In 2010, PCV13, which added serotypes 1, 3, 5, 6A, 7F, and 19A, replaced PCV7 in the Navajo Nation for children age <5 years, and routine vaccination with PCV13 was recommended for adults ≥65 years from 2015-2019. Beginning in 1989, the 23-valent Pneumococcal Polysaccharide Vaccine (PPSV23) was recommended for use among Native American individuals, given the increased incidence of IPD in this population. Over the years, and with the introduction of PCVs, recommendations for PPSV23 use evolved to focus on use in older adults and those with underlying conditions placing them at increased risk of IPD [20]. By 2019, 85-89% of children aged 2 years or less had received four doses of PCV13 [21], and substantial reductions in vaccine serotype carriage and disease were documented for additional PCV13 serotypes [22,23]. Prior to the switch to PCV13, serotype 3 was among the leading causes of IPD in the Navajo Nation in both children <5 years and adults ≥18 years of age (2001-2009: <5 years: 5.8 per 100,000; ≥18 years: 3.8 per 100,000). Thereafter, rates of serotype 3 IPD remained largely unchanged, followed by a significant increase among adults from 2015-2019 (annual rates ranging from 5 to 12 per 100,000) (Supplemental Figure 1) [24]. To understand the epidemiologic and genomic characteristics of serotype 3 epidemiology in the Navajo Nation, we analyzed genomic and epidemiological data from cases of carriage (n=61) or IPD (n=131) from 2001–2018.

## Materials and methods

### Study population and sample selection

This study included serotype 3 pneumococci isolated from cases of IPD and pneumococcal carriage identified among Indigenous individuals in the Navajo Nation in the Southwest US. IPD isolates were collected from IPD cases identified through active, laboratory-based surveillance conducted by the Johns Hopkins Center for Indigenous Health (CIH) in partnership with the Navajo Epidemiology Center [25]. Briefly, active surveillance is carried out at all clinical microbiology laboratories serving the Navajo Nation to identify cases of IPD. For each case, information on case demographics, vaccination history, clinical syndrome, and outcomes are abstracted from the medical chart. Pneumococcal isolates are sent to the Arctic Investigations Program (AIP) at the Centers for Disease Control and Prevention (CDC) in Anchorage, Alaska, for serotyping by slide agglutination, with confirmation by the Quellung reaction. Serotype 3 isolates from IPD cases identified between 2001 and 2018 were included in this analysis (n=131). In addition, serotype 3 carriage isolates from healthy individuals among three studies in the Navajo Nation were included: 1) isolates from participants aged <5 years or ≥18 years enrolled in a longitudinal household cohort study from 2006-2008 (n=31) [26]; 2) isolates from participants aged <5 years or ≥18 years enrolled in a cross-sectional study from 2010–2012 (n=16) [22]; 3) isolates from participants aged <5 years or ≥18 years enrolled in a cross-sectional study from 2015-2017 (n=14) [27,28].

All studies contributing isolates were approved by the Johns Hopkins Bloomberg School of Public Health Institutional Review Board (IRB# 2281, 9505, 11069) and the Navajo Nation Human Research Review Board (NNR-05.165, NNR-09.253, NNR-19.343).

### Bacterial genomic DNA (gDNA) isolation and whole-genome sequencing

Bacterial gDNA extraction, library preparation, whole-genome sequencing, and genome assembly was carried out as previously described [29]. Briefly, after gDNA extraction and quantification, sequencing libraries were constructed using the Illumina Nextera Flex library prep kit, which were subsequently sequenced on an Illumina MiSeq using 600 cycle V3 chemistry flow cell to produce 2 × 250 paired end reads with a target idealized coverage of 75× [29]. Post-sequencing, reads were filtered using Trimmomatic v.0.39 and quality was assessed using FastQC [30]. *De novo* genome assembly was performed using Unicycler v0.4.8 [31], and assemblies were annotated using Prokka v1.14.15 [32]. Pangenome analysis was performed using Roary v.3.12 [33], and a core genome single-nucleotide polymorphism (SNP) alignment was extracted using snp-sites v2.4.0. Maximum likelihood (ML) phylogenies were inferred with IQTREE v1.6.8 using the ASC+GTR+GAMMA substitution model with 100 bootstrap replicates [34]. SNPs in the core genome were then used to assess population structure with fastBAPS [35] and dominant lineages were identified. Abricate was used to detect the antibiotic resistance genes, φOXC141 prophage, and protein antigen variants (https://github.com/tseemann/abricate) [36] using argannot [37] and vfdb [38] as databases, respectively.

### Genomic analysis

Clade assignment was performed using the definitions previously described in Azarian *et al* [17]. Nine reference strains from the prior analysis were used in an ML phylogeny to identify: *clade Iα, clade II* and non CC180 clade. After assigning MLST [39] and clade membership, we constructed reference-based assemblies for CC180 strains (n=149) using the OXC141 reference with snippy (https://github.com/tseemann/snippy). We then compared the global CC180 dataset (n=285) from Azarian *et al.* [17] to the extant dataset from Navajo communities in the Southwest US (n=149). Gubbins was used to assess recombination, obtain a recombination-free alignment, and infer an ML phylogeny [40].

To date the emergence of *clade II*, we performed Bayesian coalescent analysis of the CC180 isolates in the extant dataset from Navajo communities (n=149). Temporal signal (i.e., linear relationship between genetic distance and sampling time in the available sequences) was assessed in TempEst v1.5.3 [41] using a starting ML tree. The *treedater* package implemented in R v3.6.0 was then used for visualization [42]. A relaxed molecular clock and Gaussian Markov random field (GMRF) demographic model [43] was run using recombination-free SNP alignments, HKY nucleotide substitution model, and ascertainment bias correction using BEAST v.10.4 [44]. Markov chain Monte Carlo (MCMC) chain length was 100 million in triplicate runs and effective sampling size (ESS) values were assessed to determine sufficient mixing using Tracer v1.6.0 [45]. To test for evidence of expansion of CC180 clades within this dataset (n=149), the dated phylogeny was used as input for CaveDive [46]. Last, we assessed differences in invasiveness, IPD case demographics, and vaccination history among CC180 clades using questionr in R version 3.6.0 (https://github.com/juba/questionr).

## Results

Characteristics of participants contributing IPD and carriage isolates are presented in Supplemental Tables 1 and 2, respectively.

### Population structure and genomic characteristics of serotype 3 carriage and IPD isolates

The serotype 3 population structure was characterized by three dominant subpopulations: *clade II* (n=90, 46.9%) and *clade Iα* (n=59, 30.7%), which fall into CC180, and a non CC180 clade (n=43, 22.4%) (Figures 1 and 2). *Clade II* could further be subdivided into two well-supported sub-clades (IIα and IIβ; Figure 2). Among the non CC180 strains, the most prevalent sequence type was ST1119 (n=26; 13.5%), isolated mainly from IPD cases among adults (n=15). In addition, we identified a novel ST16633 strain that is a single locus variant of ST433. The proportion of *clade II*-associated IPD cases increased significantly from 2001-2010 to 2011-2018 among adults (23.1% [9/39] to 71.8% [56/78]; p<0.001) but not children (27.3% [3/11] to 33.3% [1/3]; p=0.84). Over the same period, the proportion of *clade II-*associated carriage increased; this was statistically significant among children (23.3% [7/30] to 52.6% [10/19]; p=0.04) but not adults (0% [0/4] to 50.0% [4/8]; p=0.08) (Figure 1).

**Figure 1.**
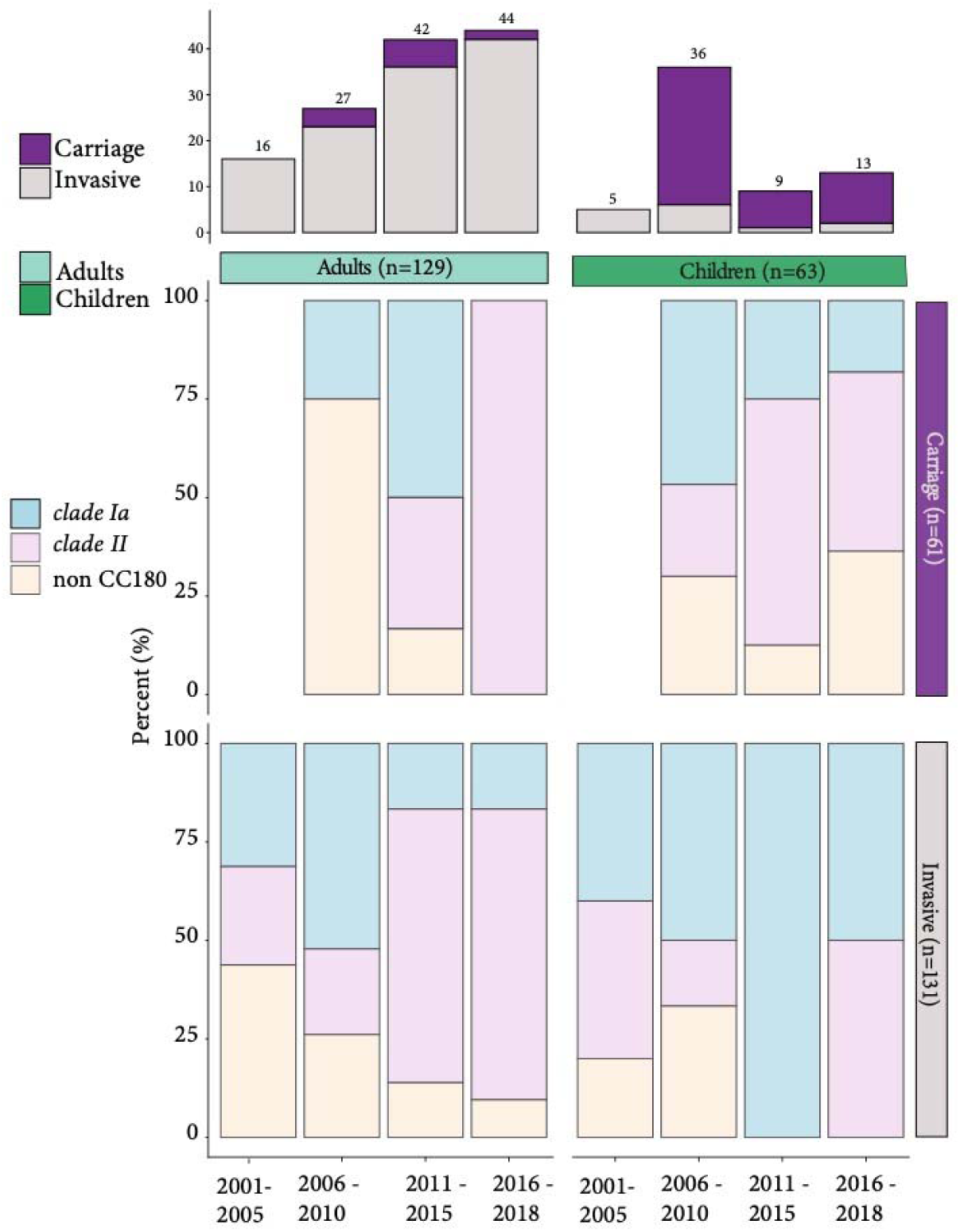
Distribution of *S. pneumoniae* serotype 3 clade membership by year, age, and source. Top panel stacked bar chart represent the distribution of carriage and invasive isolates for adults (left) and children (right) by 5-year window. Bottom panels show the proportion of carriage and invasive isolates for adults and children by 5-year window.

**Figure 2.**
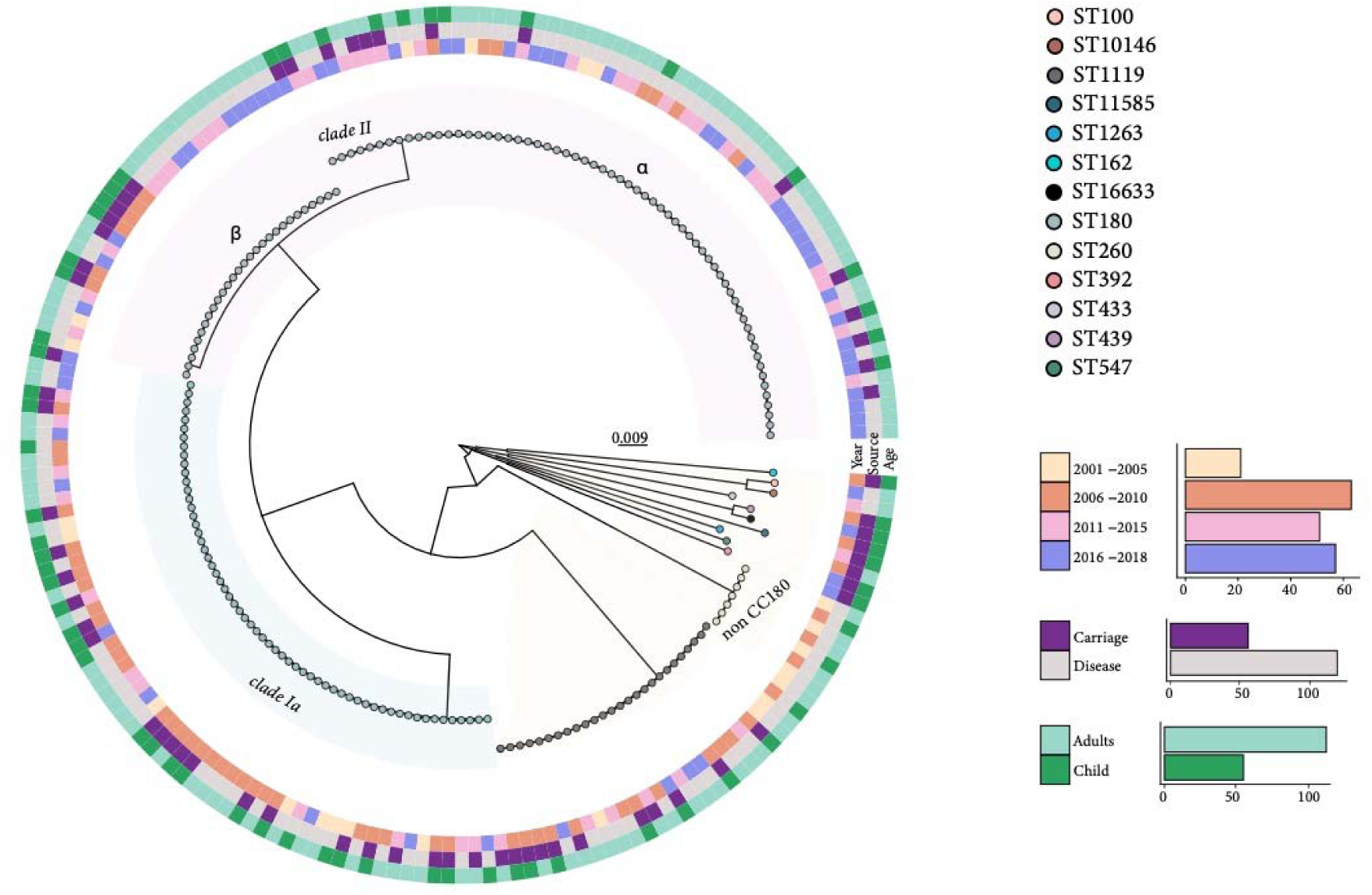
Phylogeny of *S. pneumoniae* serotype 3 isolates (n=192) showing year of collection (inner ring), collection source (invasive/carriage, middle ring) and participant age group (outside ring). The genetic distance is shown by the branch length scale bar. Tip shades indicate multilocus sequence type (MLST), and dominant subpopulations are labelled. Counts of the strains according to the different groups (collection year, collection source, age group) are shown on the bottom right.

Overall, isolates from children and adults were intermixed throughout the phylogeny, and strains typically clustered, as expected, with other isolates collected during the same time period.

Assessment of protein antigen variants in the context of population structure identified six variable protein antigens, including membrane associated protein SP2194, cell-wall anchored proteins neuraminidase A (*nanA*) and β-N-acetylhexosaminidase (*strH*), Zinc metalloprotease A (*zmpA*), and surface exposed proteins *pspC* and *pspA* (Figure 3). In particular, *pspA* varied between *clade IIα* and *IIβ,* with *clade IIβ* possessing the Family 2 variant and *clade IIα* possessing Family 1. Antibiotic resistance determinants were found to be generally lacking among CC180 strains. Only one isolate was found to possess the *Tn916* transposon harboring *erm*B and *tet*M. Further, 10 *clade II* isolates uniquely possessed tet32, which is associated with tetracycline resistance. The presence of the φOXC141 prophage was isolated to *clade I*, identified in 61.0% (36/59) of *clade I* genomes.

**Figure 3.**
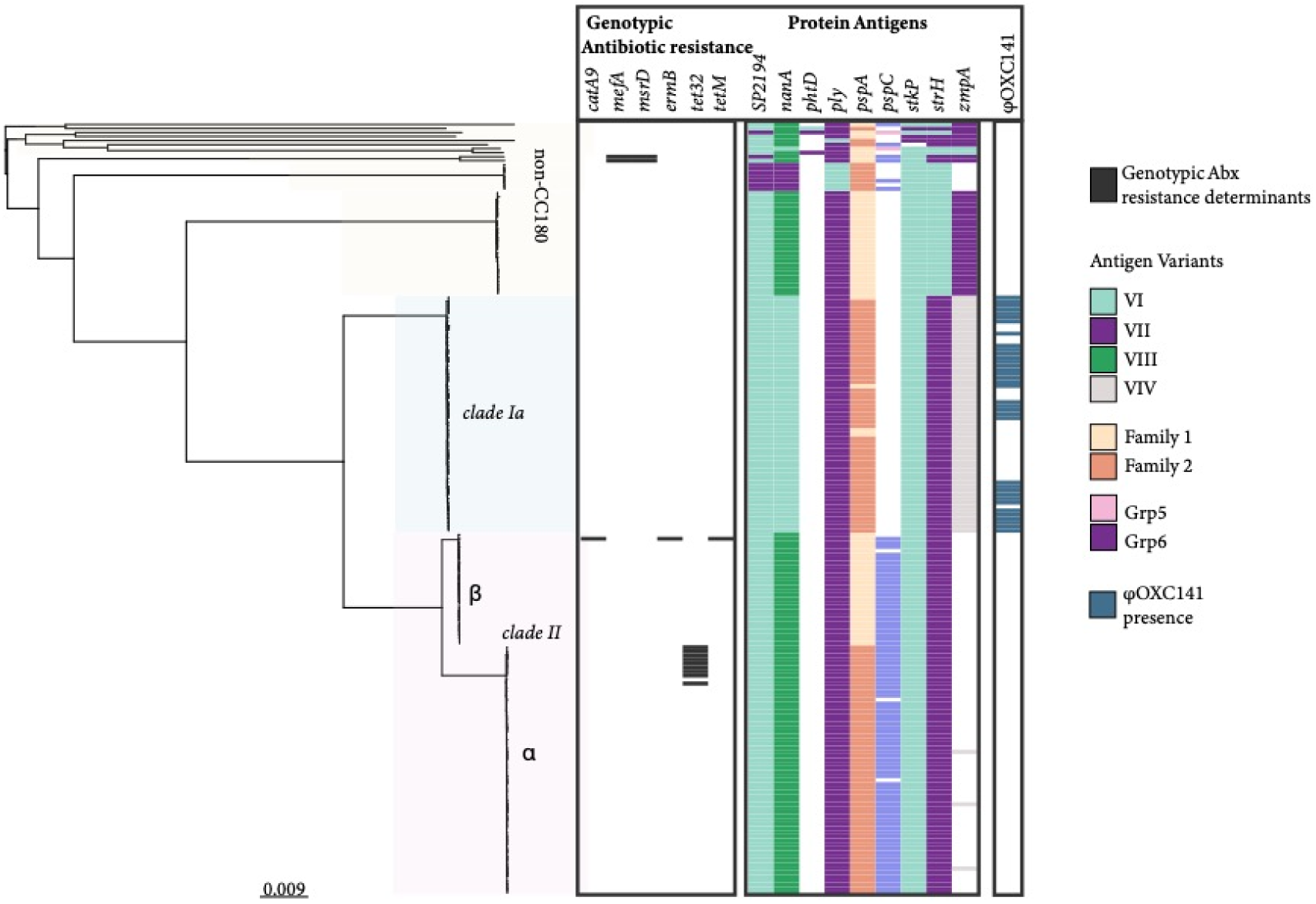
Maximum likelihood phylogeny illustrating population structure, genotypic antibiotic resistance determinants, polymorphic protein antigen variants, and φOXC141 prophage among *S. pneumoniae* serotype 3 isolates (n=192) from Navajo communities in the Southwest US. The presence of genotypic antibiotic resistance determinants is shown with a black rectangle on the heatmap. Antibiotic resistance determinants include: phenicol resistance (catA9), macrolide resistance (*mefA*, *msrD*, *ermB*) and tetracycline resistance (*tetM*, *tet32*). Corresponding protein antigen variants for SP1294, *nanA*, *phtD*, *ply*, *pspA*, and *pspC*, *strH*, and *zmpA* are illustrated on the right.

In the expanded global phylogeny of 434 CC180 genomes, which includes additional isolates from the general US population available through the Global Pneumococcal Sequencing Project database (Figure 4), *clade II* carriage and IPD isolates from this study formed two distinct clades (*clade IIα* and *clade IIβ*), whereas *clade Iα* isolates were more intermixed among global isolates suggesting a more dispersed global distribution. Investigating the ancestry of CC180 *clade IIα* and *clade IIβ* isolates from this study, we found that other isolates from the general US population were basal (i.e., closer to the root). This suggested recent independent introductions of two distinct *clade II* populations followed by expansion that we further investigated using coalescent analysis.

**Figure 4.**
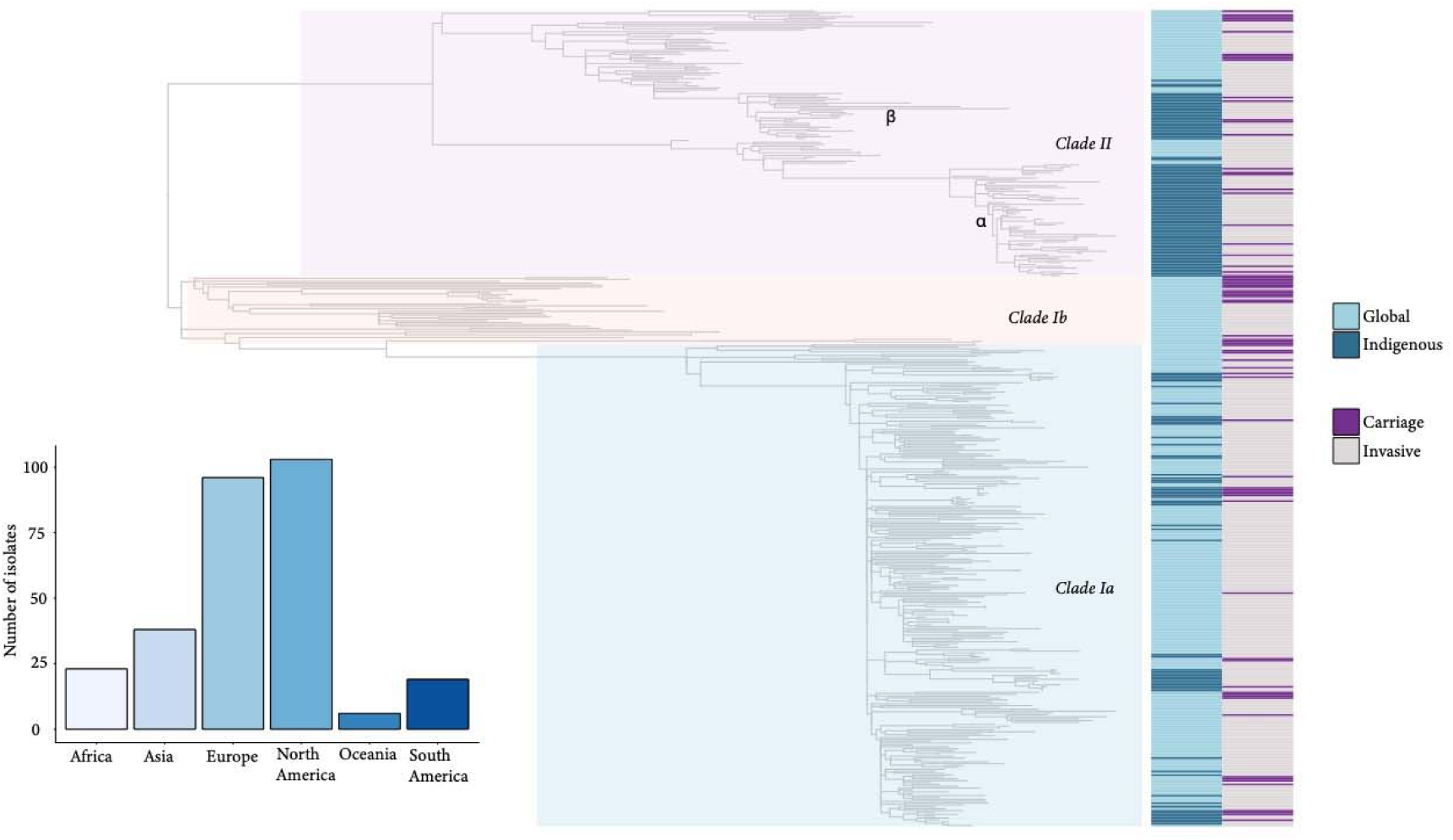
Phylogeny of *S. pneumoniae* serotype 3 CC180 isolates (n=149) with previously published global dataset (n= 285). Clade assignments are labeled on the phylogeny. The heatmap to the right indicates genomes from Global and Indigenous serotype 3 isolates and whether they were collected from carriage or disease. The bar graph on the bottom left of the figure shows the counts of serotype 3 genomes from the Global sample by geographic region of collection.

Bayesian coalescent analysis of the CC180 isolates from this study (n=149) showed a good temporal signal with significant correlation (r=0.62) between the genetic distance of each sequence to the root of the tree. The time-scaled phylogeny illustrated that *clade Iα*, whose most recent common ancestor (MRCA) was dated in the 1960s (95% Highest Posterior Density, HPD: 1960-1973), persisted as the dominant serotype 3 lineage until largely displaced by *clade II* after 2010 (Figure 5). Further examination found that the supplanting *clade II* population arose from at least two separate contemporaneous introductions in the 1990s leading to the establishment of subclades *IIα* and *IIβ*. Subclades *IIα* and *IIβ* are inferred to be distinct because their most recent common ancestor (MRCA) was dated at 1954 (95% HPD: 1940-1970), overlapping with the MRCA of *clade Iα* and suggesting their diversification predated introduction to the study population. Using CaveDive, we identified two significant population expansion events (clades highlighted in light blue): one following the initial introduction of *clade Iα* in 1969 (95% HPD: 1966-1974) and the other involving *clade IIα* in 2000 (95% HPD: 1998-2001). While *clade Iα* showed a more equal distribution of carriage and IPD isolates, *clade II*, and *IIα* in particular, predominantly included IPD isolates. Even though these IPD isolates in clade II*α* were in a slightly greater proportion of adults with IPD as compared to clade II*β*, the association was weak (p=0.075).

**Figure 5.**
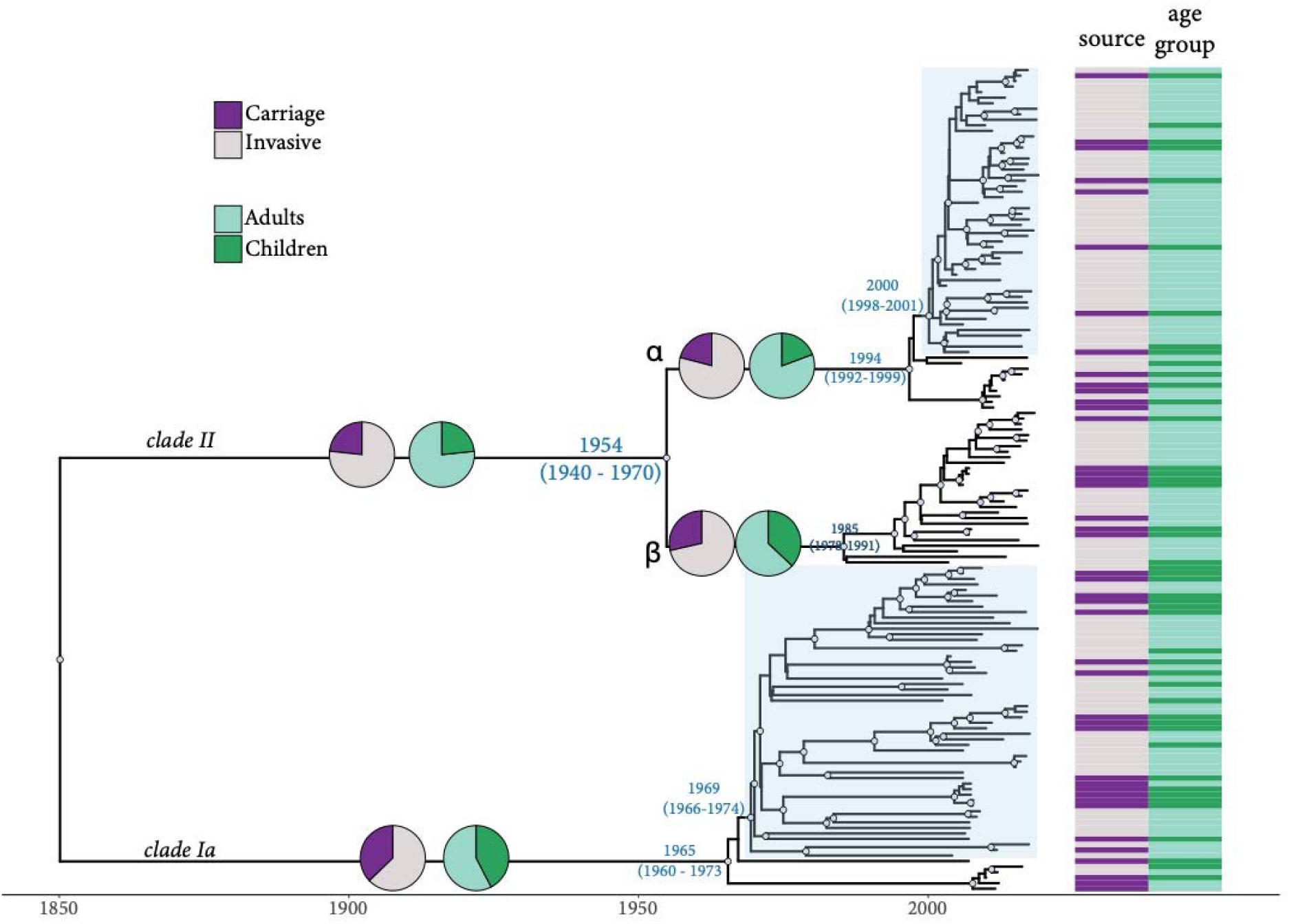
*S. pneumoniae* serotype 3 CC180 isolates dated phylogeny (n=149). Taxa are annotated according to isolation source (invasive/carriage) and age group. A white circle along the branch indicates a higher posterior probability (pp>0.8). The pie charts on the main branches show the proportion of samples by isolation source (invasive/carriage) and age group. Clades where an expansion occurred are highlighted in light blue.

### Vaccination History

The three children with IPD in the PCV13-era all had received PCV13; one case was associated with *clade II* and two were associated with *clade I*_α_. In the pre-PCV13 era, 32 IPD cases occurred among adults with vaccine history available, including 11 (34%) who had received PPSV23. Among cases associated with *clade II*, *clade 1*_α_, and non CC180, 29% (n=2/7), 38% (n=5/13), and 33% (n=4/12) had received PPSV23, respectively (p=0.90). In the PCV13-era, 67 IPD cases occurred among adults with vaccine history available, including 49 (73%) who had received at least one dose of either PPSV23 or PCV13. Among cases associated with *clade II*, *clade 1*_α_, and non CC180, 73% (n=35/48), 64% (n=7/11), and 87% (n=7/8) had received PPSV23, respectively (p=0.51).

## Discussion

*S. pneumoniae* serotype 3 CC180 is the most prevalent lineage of serotype 3 worldwide [17], and serotype 3 continues to contribute significantly to the global burden of IPD. The emergence of *clade II* represented a profound shift in the CC180 population structure. Recent observational studies have documented that serotype 3, now composed largely of isolates belonging to *clade II*, is responsible for an increasing proportion of IPD [17,18,47,48]. Using data from active surveillance and observational studies spanning almost 20 years and encompassing the introduction of PCV7 and PCV13 [17,49,50], we documented that serotype 3 strains belonging to *clade II* were present at the earliest time point, predating the introduction of PCV7. Subsequently, *clade II* steadily rose in prevalence to become the dominant cause of serotype 3 associated IPD despite the continued co-circulation of *clade Iα* and non-CC180 strains in carriage.

Our findings elucidate the demographic history of *clade II*’s emergence and further resolve the population structure. Serotype 3 strains belonging to non-CC180 lineages persisted at background levels, making up almost a quarter of all serotype 3 strains studied. The most common non-CC180 strains belonged to ST260 and ST1119, the latter of which was first identified among children in Mexico and appears to have a limited geographic distribution [51]. Here we found ST1119 equally distributed in carriage and disease with decreasing prevalence after the introduction of PCV13. Within CC180, we observed the previously defined *clade Iα* and *clade II,* but found *clade II* was further bifurcated into two distinct subclades, designated *IIα* and *IIβ*, that have contemporaneously circulated. In the context of a previously described global sample of CC180, these subclades appeared to represent two distinct introductions and not a recent divergence event [17]. Epidemiological differences between the subclades remain unclear. Population dynamic analysis suggested that *clade IIα* experienced significant population expansion, and while *clade IIα* strains were found in a slightly greater proportion of adults with IPD as compared to *clade IIβ*, this association was weak. Nevertheless, the results suggest that the expansion of *clade II* may be associated with the increase in IPD observed among adults in this population beginning in 2015.

Several factors may have contributed to the shift in the serotype 3 population including increased antibiotic resistance, differential vaccine effectiveness, divergent antigenic profile, and changes in carriage duration and invasiveness. We previously suggested that the presence of the Tn*916*-like transposon conferring multidrug resistance as well as variation in pneumococcal surface protein antigens may have contributed to the rise of *clade II* [17]. Here we found that *clade II* strains generally lacked antibiotic resistance determinants, suggesting that resistance may only be a contributing cause in regions with high antibiotic pressure and circulation of serotype 3 strains harboring the Tn*916*-like transposon. We additionally observed diversity among protein antigens including pneumococcal surface protein A (PspA), a virulence factor that interferes with complement opsonization and a putative target for non-capsular vaccines. Previously, *clades I* and *II* of CC180 were found to possess different families of PspA. This suggested that the emergent *clade II* may have had an evolutionary advantage due to population level immunity to the *clade I* protein antigen profile, as carriage and disease generate family-specific PspA antibodies [52]. However, here we found additional variation in PspA between *clade II* subclades, emphasizing two distinct evolutionary histories prior to introduction in the current study population and further obscuring the role of protein antigen variation in the CC180 shift.

The temporality of *clade II*’s increase and corresponding global increase in serotype 3 IPD in relation to the introduction of PCV13 posits a putative association. Surveillance data from countries that introduced PCV13 and PCV10 (which lacks the serotype 3 component) suggests at least some direct and indirect protection against serotype 3 from PCV13 [53–55]. While we observed serotype 3 IPD among fully vaccinated children and adults in this study, our design did not permit assessment of vaccine effectiveness nor differential incidence rates of *clade I* and *clade II* disease among vaccinated and unvaccinated individuals. The question of why an increase was not observed earlier when PCV7 was first introduced has been posed [56]. Indeed, this was a period when non-vaccine serotype replacement was broadly identified [57]. One putative explanation may be that *clade II* was more apt at “filling the hole” after the removal of vaccine serotypes through previously described negative frequency dependent selection (NFDS) dynamics and that this did not occur until the combined impact of PCV7 and PCV13 [58].

However, as both clades belong to the same CC, they likely possess similar fitness in the post-PCV landscape due to minimal differences in pangenome content, which are suggested to be the loci under selection in NFDS models. Alternatively, while *clade II* strains were found to have greater *in vitro* susceptibility than *clade I* to neutrophil-mediated opsonophagocytic killing in the presence of antisera against both PCV13 and serotype 3 polysaccharide, unmeasured differential vaccine effectiveness among clades may still contribute to varying indirect vaccine protection. At this time, however, there are no data to support this, and it remains difficult to conduct serotype 3 vaccine effectiveness studies due to its relatively low carriage prevalence compared to other pneumococcal serotypes. Taken together, the relationship between vaccine use and the emergence of *clade II* remains largely unclear. One consideration worth noting is that the timescale at which lineages or “variants” rise and fall remains an unresolved question. While initiatives like the Global Pneumococcal Sequencing project have generated over 20,000 pneumococcal genomes [59], the sample could largely be considered cross-sectional in evolutionary time. Therefore, the emergence of *clade II* and similar evolutionarily successful lineages could be stochastic and potentially common if we were to examine pneumococcal population genomics over decades.

The shift in the serotype 3 population remains an intriguing epidemiological and evolutionary question with significant implications for disease control and vaccine development. Likely, subtle genomic differences between *clade I* and *clade II* strains have resulted in phenotypic variation and subsequent observed epidemiologic trends. For example, there is emerging evidence that *clade II* demonstrates decreased invasiveness, longer carriage duration, and higher carriage density in a mouse model compared to *clade I* [60]. As carriage duration has been shown to be positively correlated with recombination rate [61], this finding would be consistent with our previous observation that *clade II* has accumulated more diversity through recombination (i.e., a higher *r/m* or ratio of SNPs introduced through recombination to the number introduced through mutation) than *clade I* [17]. Competence is also known to be closely linked with virulence [62]. One possible cause for a longer carriage duration and higher carriage density could be increased resistance to antibody mediated agglutination resulting in decreased nasopharyngeal clearance [63]. Taken together, increased carriage duration and density could explain the increase in *clade II* associated IPD as both properties would result in increased transmission and higher prevalence. Even if strains belonging to *clade II* are less invasive, a significant increase in carriage prevalence especially among unvaccinated adults could result in a considerable increase in IPD rates. More work is needed to assess epidemiological differences such as carriage prevalence and duration as well as the associated genomic variation that has precipitated such changes.

This study was not without limitations. While IPD cases were obtained through ongoing active disease surveillance, carriage data were from repeated cross-sectional studies largely focused on healthy children ≤5 years. For this reason, we were unable to directly compare the increase in serotype 3 IPD to the carriage prevalence among adults. Further, carriage isolates were only available starting in 2006, which limited our ability to identify *clade II* earlier in the study period.

Overall, we elucidated differences in epidemiologic and genomic characteristics underlying the shift in serotype 3 epidemiology toward *clade II* in the Navajo Nation in the Southwest US. This shift has been observed in the general US population and globally where CC180 circulates. We additionally highlight the benefits of pathogen genomic investigation and detailed characterization of pneumococcal populations in the context of routine vaccine use. Continued monitoring of the distribution of the serotype 3 population in relation to carriage and IPD is essential. As PCVs with varying valency are being considered in different geographic settings, genomic surveys of pneumococcal populations should be considered before and after vaccine introduction to assess changes in serotype distribution as well as lineage composition.

## Data Availability

All data produced in the present study are available upon reasonable request to the authors

## Acknowledgements

Special thanks to the participants from each study, study staff of the Center for Indigenous Health and Navajo Epidemiology Center, the Navajo Nation Human Research Review Board and the JHSPH IRB. The content is solely the responsibility of the authors and does not necessarily represent the views of the US Centers for Disease Control and Prevention.

