## Supplemental Material for "*Streptococcus pneumoniae* serotype 3 population structure in the era of conjugate vaccines, 2001-2018"

**Supplemental Table 1.** Characteristics of cases with serotype 3 invasive pneumococcal disease, Navajo Nation, 2001-2018

|  | **Total**  **(n=131)** | **2001-2005**  **(n=21)** | **2006-2010**  **(n=29)** | **2011-2015**  **(n=37)** | **2016-2018**  **(n=44)** | **p-value** |
| --- | --- | --- | --- | --- | --- | --- |
| Sex – female | 55 (42.0%) | 10 (47.6%) | 12 (41.4%) | 15 (40.5%) | 18 (40.9%) | 1 |
| Age group (years) |  |  |  |  |  |  |
| <5 | 14 (10.7%) | 5 (23.8%) | 6 (20.7%) | 1 (2.7%) | 2 (4.5%) | 0.008 |
| 5-17 | 0 (0%) | 0 (0%) | 0 (0%) | 0 (0%) | 0 (0%) |  |
| 18-49 | 25 (19.1%) | 3 (14.3%) | 8 (27.6%) | 7 (18.9%) | 7 (15.9%) |  |
| 50-64 | 46 (35.1%) | 4 (19.0%) | 12 (41.4%) | 12 (32.4%) | 18 (40.9%) |  |
| ≥65 | 46 (35.1%) | 9 (42.9%) | 3 (10.3%) | 17 (45.9%) | 17 (38.6%) |  |
| Source of isolate |  |  |  |  |  |  |
| Blood | 126 (96.2%) | 21 (100.0%) | 28 (96.6%) | 35 (94.6%) | 42 (95.5%) | 1 |
| Cerebrospinal fluid | 2 (1.5%) | 0 (0%) | 0 (0%) | 1 (2.7%) | 1 (2.3%) |  |
| Pleural fluid | 3 (2.3%) | 0 (0%) | 1 (3.4%) | 1 (2.7%) | 1 (2.3%) |  |
| Clinical syndrome^a^ |  |  |  |  |  |  |
| Pneumonia | 112 (85.5%) | 15 (71.4%) | 25 (86.2%) | 32 (86.5%) | 40 (90.9%) | 0.248 |
| Meningitis^b^ | 5 (3.8%) | 1 (4.8%) | 0 (0%) | 2 (5.4%) | 2 (4.5%) | 0.665 |
| Non pneumonia/ meningitis | 17 (13.0%) | 5 (23.8%) | 4 (13.8%) | 5 (13.5%) | 3 (6.8%) | 0.281 |
| PCV13 doses among children <5 years in 2011-2018 (n=3) |  |  |  |  |  | 1 |
| ≥3 | 3 (100%) | n/a | n/a | 1 (100.0%) | 2 (100.0%) |  |
| PPV23 and PCV13 doses among adults ≥50 years (n=92) |  |  |  |  |  | 0.001 |
| 0 | 25 (27.2%) | 4 (30.8%) | 8 (53.3%) | 6 (20.7%) | 7 (20.0%) |  |
| ≥1 PPSV23 only | 44 (27.8%) | 8 (61.5%) | 3 (20.0%) | 20 (69.0%) | 13 (37.1%) |  |
| ≥1 PCV13 only | 1 (1.1%) | n/a | n/a | 0 (0%) | 1 (2.9%) |  |
| ≥1 PPSV23 and PCV13 | 11 (12.0%) | n/a | n/a | 0 (0%) | 11 (31.4%) |  |
| Unknown | 11 (12.0%) | 1 (7.7%) | 4 (26.7%) | 3 (10.3%) | 3 (8.6%) |  |
| Any underlying medical conditions^c^ | 100 (76.3%) | 12 (57.1%) | 21 (72.4%) | 26 (70.3%) | 41 (93.2%) | 0.003 |
| Hospitalized^d^ | 125 (97.7%) | 20 (100%) | 26 (89.7%) | 37 (100%) | 42 (100%) | 0.014 |
| Outcome – died^e^ | 15 (12.5%) | 5 (25.0%) | 2 (7.4%) | 2 (6.1%) | 6 (14.3%) | 0.190 |

n/a: not applicable; PCV: pneumococcal conjugate vaccine; PPSV: pneumococcal polysaccharide vaccine

^a^ Individuals may have been diagnosed with more than one syndrome

^b^ Three cases presenting with meningitis had blood but not cerebrospinal fluid collected for culture

^c^ Underlying conditions assessed by medical chart review and restricted to those identified by the Advisory Committee on Immunization Practices (ACIP) as warranting administration of pneumococcal vaccines [64]

^d^ Admission status missing for 1 case in 2001-2005

^e^ Outcome missing for 11 cases (1 in 2001-2005; 2 in 2006-2010; 4 in 2011-2015; 4 in 2016-2018)

**Supplemental Table 2.** Characteristics of participants with serotype 3 carriage, Navajo Nation, 2006-2018

|  | **Total**  **(n=61)** | **2006-2010**  **(n=34)** | **2011-2015**  **(n=14)** | **2016-2018**  **(n=13)** | **p-value** |
| --- | --- | --- | --- | --- | --- |
| Sex – female | 28 (45.9%) | 18 (52.9%) | 5 (35.7%) | 5 (38.5%) | 0.510 |
| Age group (years) |  |  |  |  | 0.070 |
| <5 | 43 (70.5%) | 26 (76.5%) | 6 (42.9%) | 11 (84.6%) |  |
| 5-17 | 6 (9.8%) | 4 (11.8%) | 2 (14.3%) | 0 (0%) |  |
| 18-49 | 9 (14.8%) | 4 (11.8%) | 4 (28.6%) | 1 (7.7%) |  |
| 50-64 | 1 (1.6%) | 0 (0%) | 1 (7.1%) | 0 (0%) |  |
| ≥65 | 2 (3.3%) | 0 (0%) | 1 (8.1%) | 1 (7.7%) |  |
| PCV13 doses among children <5 years (n=17) |  |  |  |  | 0.052 |
| 0 | 2 (11.8%) | n/a | 1 (16.7%) | 1 (9.1%) |  |
| 1-2 | 5 (31.3%) | n/a | 3 (50.0%) | 2 (18.2%) |  |
| ≥3 | 10 (62.5%) | n/a | 2 (33.3%) | 8 (72.7%) |  |
| PPV23 and PCV13 doses among adults ≥50 years (n=3) |  |  |  |  | 1 |
| 0 | 2 (66.7%) | 0 (0%) | 1 (50.0%) | 1 (100.0%) |  |
| ≥1 PPSV23 only | 1 (33.3%) | 0 (0%) | 1 (50.0%) | 0 (0%) |  |
| ≥1 PCV13 only | 0 (0%) | n/a | 0 (0%) | 0 (0%) |  |
| ≥1 PPSV23 and PCV13 | 0 (0%) | n/a | 0 (0%) | 0 (0%) |  |

n/a: not applicable; PCV: pneumococcal conjugate vaccine; PPSV: pneumococcal polysaccharide vaccine

**Supplemental Figure 1.** Incidence of serotype 3 invasive pneumococcal disease in adults (top) and children (bottom), Navajo Nation, 2000-2019

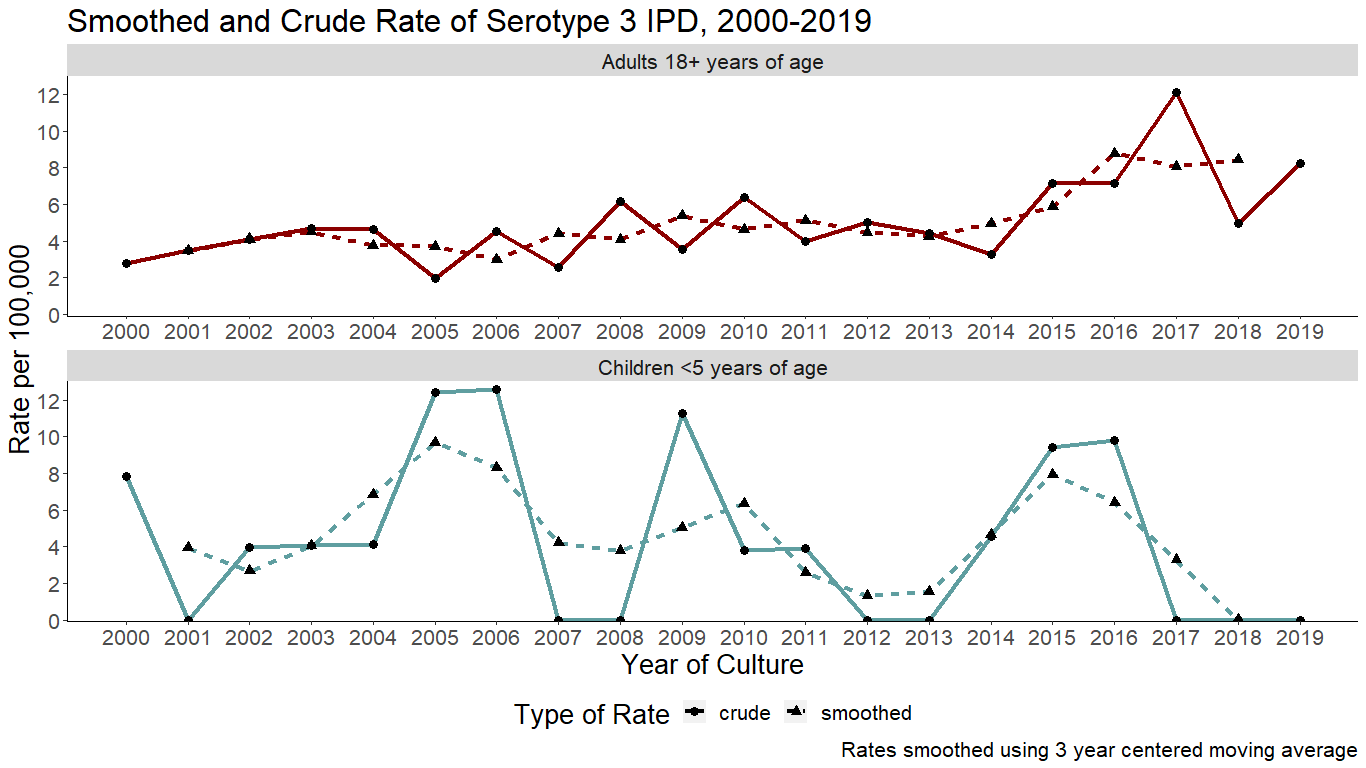
